# Machine learning based prediction of high school student mental health

**DOI:** 10.1101/2025.08.07.25333218

**Authors:** Wesley Lo, Senbao Lu, Dmitry Korkin, Angela C. Incollingo Rodriguez, Lourah M. Kelly, Jean A. King, Benjamin C. Nephew

## Abstract

Recent increases in the prevalence rates of anxiety, depression, and suicidal ideation, especially in student populations, present an urgent need to develop targeted diagnostic and treatment tools. Given the substantial evidence of variation in mental health symptomatology, efforts to develop prevention and intervention strategies may benefit from machine learning based investigations of individual and group variability in predictors of anxiety, depression, and suicidal ideation. The present study investigated the use of random forest classifiers (RFC) in predicting anxiety and depression screening scores and suicidal ideation in 9^th^ grade students from a Massachusetts public school district (n = 274). Our highly accurate (80-95%) predictive analyses identified a strong inverse relationship between academic performance and depression and anxiety and substantial gender and race based variation in specific academic and symptom level predictors of anxiety, depression, and suicidal ideation. These findings illustrate the need for personalized clinically relevant data collection and analyses to inform mental health programming, preventative measures, and interventions.

## INTRODUCTION

As anxiety, depression, and suicide become more prevalent among adolescents and young adults around the globe, there is a growing need for the accurate identification of at-risk individuals to target and inform treatment. The 2023 CDC Youth Behavior Risk Survey, which highlights youth trends over the decade from 2013 to 2023, reported that 4 out of 10 high school students reported feeling “persistently sad or hopeless” over the past year, with rates varying by gender and race. Compared to male peers, female students are almost twice as likely to feel persistent feelings of sadness or hopelessness (53% vs. 28%). A recent meta-analysis of 350,000 college students from 373 campuses across 2013-2021 found that although >60% of students met criteria for one or more mental health problems, a nearly 50% increase from 2013, students of color had the lowest rates of mental health service utilization (Lipson et al., 2022; Verlenden, 2024). Although these increases have been exacerbated by the COVID-19 pandemic (Hawes et al., 2022), rates were increasing prior to COVID and potentially exacerbated by the COVID-19 pandemic. These trends in increased prevalence support the need for improved prediction, diagnosis and treatment of mental illness, especially in younger populations. Harmonizing across various potential predictors can be challenging, especially when considering multi-modal data.

Predictive Machine Learning (ML) and Deep Learning (DL) algorithms are ideal in this context and have been successfully deployed in mental health research (Iyortsuun et al., 2023; Su et al., 2020; Su et al., 2022). Machine learning can lead to more targeted and effective interventions (Shatte et al., 2019). Several recent studies showcase high predictive performance across diverse data conditions, ranging from biomarkers and genomics to psychiatric treatment responses (Abdul Mumin et al., 2023; Ehiabhi and Wang, 2023; Kautzky et al., 2017).

Investigations focused specifically on youth mental health have demonstrated accurate prediction of mental health outcomes (Sajjadian et al., 2023; Tate et al., 2020). Despite these encouraging developments, it is still extremely rare for high schools or colleges to collect the type of data needed to take advantage of the recent advances in applying ML and DL to mental health objectives. The few that do are unlikely to have the resources necessary to conduct ML and DL based analyses and take advantage of the associated findings.

In response to the growing concerns about student mental health, some U.S. school districts have begun to collect clinical mental health screening data to assist in identifying at-risk students. The primary objective of the present study was to predict anxiety and depression outcomes using demographic, academic and individual mental health screening tool questions in a Massachusetts public school 9^th^ grade class using the PHQ-9 for depression and GAD-7 for anxiety. These screening tools are efficient and selective diagnostic tools for anxiety, depression, and suicidal ideation (Löwe et al., 2008a; Löwe et al., 2008b; Manea et al., 2015) and provide details of specific mental health symptomology including anhedonia, hopelessness, and fatigue. We used the Random Forest Classifier (RFC) to predict overall PHQ9 and GAD7 scores, which allowed for the determination of which questions were most responsible for predicting mental health outcomes. While the immediate value of using these screening tools to identify at-risk students is critically important, the additional predictive analyses in the present study provide additional valuable input on overall and group specific patterns which can inform academic and mental health programming, prevention, and intervention effects.

Based on initial observations of gender and race specific differences in predictors of anxiety, depression, and suicidal ideation outcomes, we assessed RFC performance by gender and race.

## METHODS

### Data Availability and Summary

The deidentified dataset was provided by the high school administration of a Massachusetts public school district through a data use agreement and is not publicly accessible. Students, and parents on their behalf, were given the option to opt out of being included in the dataset and study, and 9 out of 329 opted out. The dataset contained information for 9^th^ grade students (class of 2025, collected in October 2022). The GAD-7 and PHQ-9 are widely used self-reporting questionnaires for screening anxiety and depression, respectively, modeled after the Diagnostic and Statistical Manual of Mental Disorders, Fourth Edition (DSM-IV)(Guze, 1995). The questions and short descriptors are listed in Table 1. Tables 4-6 present GAD-7, PHQ-9 and endorsement of PHQ-9 question 9 distributions.

**Table 1.**
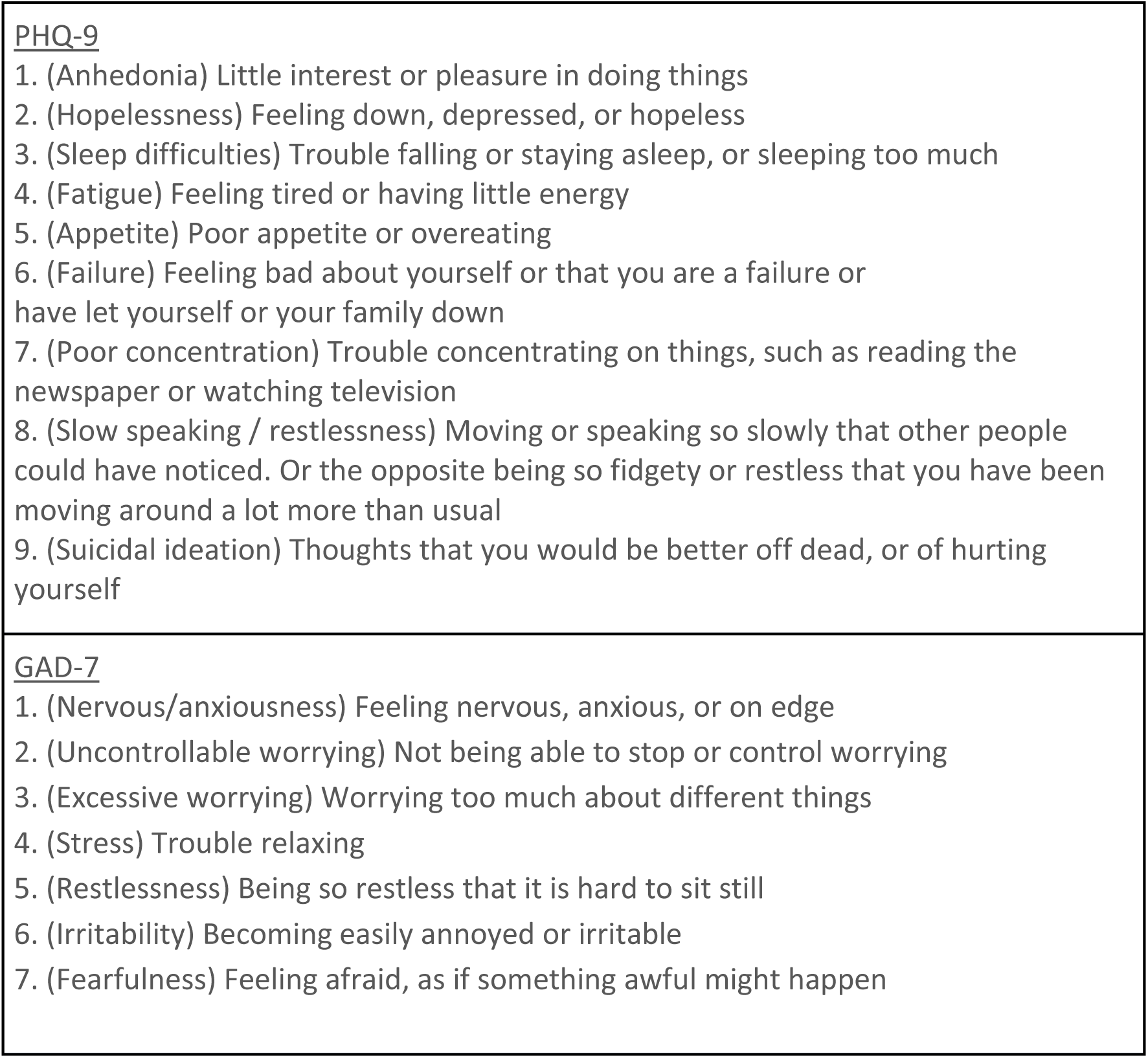
Labeled PHQ-9 and GAD-7 questions (Kroenke et al., 2001; Spitzer et al., 2006)

**Table 2.** PHQ-9 symptoms scoring (Kroenke et al., 2001)

| Score | Symptom Severity | Comments |
| --- | --- | --- |
| 0-4 | Minimal or none | Monitor; treatment may not be needed |
| 5-9 | Mild | Consider symptom duration, functional impairment to determine whether treatment is necessary |
| 10-14 | Moderate |  |
| 15-19 | Moderately severe | Active treatment with psychotherapy, medication, or a combination of both is warranted. |
| 20-27 | Severe |  |

**Table 3.** GAD-7 symptoms scoring(Spitzer et al., 2006)

| Score | Symptom Severity | Comments |
| --- | --- | --- |
| 0-5 | None | Monitor; treatment may not be needed |
| 5-9 | Minimal |  |
| 10-14 | Moderate | Possibly clinically significant |
| >15 | Severe | Active treatment is possibly warranted |

**Table 4.** Percents and n’s of GAD-7 score distribution.

| Gender | Female |  | Male |  | All |
| --- | --- | --- | --- | --- | --- |
| Race / Ethnicity | Asian | White | Asian | White |  |
| **None (0-4)** | 8.03% (22) | 9.85% (27) | 4.74% (13) | 8.76% (24) | 31.39% (86) |
| **Minimal (5-9)** | 8.03% (22) | 10.95% (30) | 11.68% (32) | 16.79% (46) | 47.45% (130) |
| **Moderate (10-14)** | 3.28% (9) | 7.30% (20) | 1.09% (3) | 2.19% (6) | 13.87% (38) |
| **Severe (>15)** | 1.09% (3) | 4.38% (12) | 0.36% (1) | 1.46% (4) | 7.30% (20) |
| **All** | 20.44% (56) | 32.48% (89) | 17.88% (49) | 29.20% (80) | 100.00% (274) |

**Table 5.** Percents and n’s of PHQ-9 score distribution.

| Gender | Female |  | Male |  | All |
| --- | --- | --- | --- | --- | --- |
| Race / Ethnicity | Asian | White | Asian | White |  |
| **None-Minimal (0-4)** | 16.06% (44) | 21.90% (60) | 16.06% (44) | 25.91% (71) | 79.93% (219) |
| **Mild (5-9)** | 1.82% (5) | 5.47% (15) | 1.09% (3) | 2.19% (6) | 10.58% (29) |
| **Moderate (10-14)** | 2.55% (7) | 3.28% (9) | 0.73% (2) | 0.73% (2) | 7.30% (20) |
| **Moderately Severe & Severe (>15)** | 0.00% (0) | 1.82% (5) | 0.00% (0) | 0.36% (1) | 2.19% (6) |
| **All** | 20.44% (56) | 32.48% (89) | 17.88% (49) | 29.20% (80) | 100.00% (274) |

**Table 6.** Percents and n’s of Q9 endorsement score distribution.

| Gender | Female |  | Male |  | All |
| --- | --- | --- | --- | --- | --- |
| Race / Ethnicity | Asian | White | Asian | White |  |
| None | 17.52%<br>(48) | 28.10%<br>(77) | 16.06%<br>(44) | 26.28%<br>(72) | 87.96% (241) |
| Endorsed | 2.92% (8) | 4.38% (12) | 1.82% (5) | 2.92% (8) | 12.04% (33) |
| All | 20.44%<br>(56) | 32.48%<br>(89) | 17.88%<br>(49) | 29.20%<br>(80) | 100.00%<br>(274) |

### Scoring PHQ-9 and GAD-7

Responses to each question for both measures are scored on a scale of 0 to 3, with higher scores indicating greater severity of symptoms (Kroenke et al., 2001; Spitzer et al., 2006). For the PHQ-9, an individual’s summary score is obtained by summing all question responses, thus ranging from 0-27. Specific cutoffs indicate increasing severity: minimal (0–4), mild (5–9), moderate (10–14), moderately-severe (15–19), and severe (20–27, table 2). The GAD-7 summary score is the sum of all question responses ranging up to 21 with the following cutoffs: minimal (0–4), mild (5–9), moderate (10–14), and severe (15–21, table 3). Q9 endorsement in the present study refers to any score greater than 0 (Kroenke et al., 2001).

The following features were also included in the analyses: academic performance (year-end GPA weighted or unweighted), educational statuses (special education (SPED), English-language learner (ELL), and 504 plan), and race / ethnicity and gender. There were 274 students in the filtered dataset.

### Data Analysis Tools

All data analysis and visualizations were provided from Python and its libraries. Pandas was used to clean the dataset and host it as a dataframe for analysis (McKinney, 2010). Sklearn was used for binary classification models (random forest, logistic regression) (Pedregosa et al., 2011). Seaborn and Matplotlib were used to make heatmaps and plots (Hunter, 2007; Waskom, 2021). Analysis of variance (ANOVA) was performed using Pingouin (Vallat, 2018).

### Preprocessing

We filtered the dataset using Pandas. The numbers of Black and gender non-conforming students were too low (<10 students) to be analyzed in the present analyses, which was then restricted to Asian, White, male, and female groups. We also removed rows with missing values. This left 274 students in the year-end dataset.

### Feature Sets

Two types of feature sets were used in our RFC analyses. The summary score analyses used educational indicators (SPED, ELL, 504 plan status), academic performance (GPA), and comorbid mental health scores (e.g., GAD-7 total score and PHQ-9 item 9 suicidal ideation endorsement when predicting PHQ-9). The question level analyses included individual responses to both PHQ-9 and GAD-7 questions alongside the educational and academic features. In all analyses, only the target variable was binarized while predictor variables retained their original values.

### ANOVA

Two-way ANOVA was performed to discern both main effects and specific interaction effects between Race / Ethnicity and Gender across individual GAD-7 and PHQ-9 questions and full summary scores. Pingouin’s “ANOVA” function was used for this, where gender and race were input as a list in the between keyword and each feature of interest as dv (data value).

### Random Forest Classification & Feature Contributions

For the prediction of mental health status in the WHS mental health dataset, we settled on using the random forest classifier (RFC), an ensemble learning method that leverages decision trees to make binary classifications. One benefit of using RFC over other algorithms is the interpretability through feature contributions. Random forest as a decision-tree-based model can capture causal relationships between features in its prediction process, through feature contributions. These results sum to 1 and represent how well RFC’s decision trees divide the data into positive and negative classes via Gini importance. In prediction of each target, feature contributions for each feature were averaged across all five models. The top two features of this aggregate were included in the results section. This analysis was run using instances of Sklearn’s RandomForestClassifier class. The following grid was used to train these models:

’n_estimators’: [250, 500, 1000, 1250]

’max_depth’: [1, 5, 10]

’min_samples_split’: [2, 5, 10]

’min_samples_leaf’: [1, 2, 4]

### Predictive Analysis Pipeline

For predictive analyses, we stratified by gender, race / ethnicity, and both, yielding 8 groups. Stratification was done by indexing the dataframe using pandas. In each group, 6 total analyses were performed; predictions on the three targets (GAD-7 predictions, PHQ-9 predictions, and Q9 endorsement predictions) for question level and summary score level features.

We sought to avoid two problems in the formation of test and train sets: (1) keep the proportion of mental health targets the same in both test/train sets (2) re-randomize the set to capture the full population. Problem 1 was addressed using stratification, which encourages the test/train/split shuffling process towards proportions of outcome variables in splits representative of the entire dataset. For this, we used Sklearn’s StratifiedShuffleSplit function, with n_splits=5 and test-size=0.3. Problem 2 was addressed by generating five of these models or “splits’’ then averaging the results, notated by the outer shell in figure 5. Five-fold cross-validation was performed on the training set for each split. This process is outlined above in Figure 1. This whole procedure was performed once on the whole population, and also once per each of the 8 groups stratified by race / ethnicity, gender, or both.

**Figure 1.**
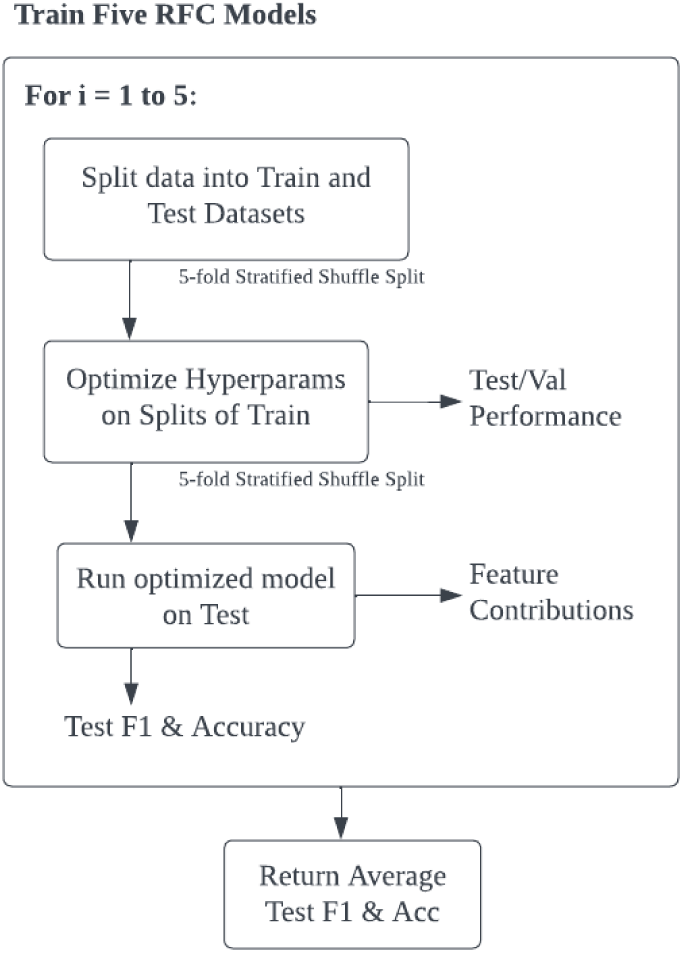
Pipeline for training Random Forest Classifiers (RFC) in given group

### GAD-7 and PHQ-9 Binarization and Group Analyses

Mental health scores were binarized according to clinical thresholds. For both GAD-7 and PHQ-9, scores ≥10 were classified as high risk (1) and scores <10 as low risk (0), aligning with validated cutoffs for moderate or greater symptom severity. For Q9 endorsement, which screens for suicidal ideation, any non-zero score (>0) was classified as high risk.

## RESULTS

Overall and group specific prevalence rates are displayed in tables 4-6. Moderate to severe anxiety was present in 21% of the students, moderate to severe depression was identified in 10%, and 12% of the students reported suicidal ideation through the endorsement of PHQ question 9. Rates of all outcomes were higher in female and White populations.

### ANOVA of GAD-7 and PHQ-9 questionnaires

Two-way ANOVA was performed for binarized survey data of anxiety (GAD-7), depression (PHQ-9), and suicidal ideation (PHQ-9 Question 9). For GAD-7 score, there were significant differences across gender and race/ethnicity. For gender, female students showed higher anxiety levels compared to male students (F = 26.028, p < 0.01), and White students had higher anxiety levels compared to Asian students (F = 4.902, p =.027). There was no significant interaction effect observed between gender and race/ethnicity (F = 0.129, p = 0.7).

When predicting PHQ-9 score, significant differences were only observed across gender.

Moderate or severe depression scores were more common in female students than male students along gender (F = 17.744, p <.001). No significant differences observed in race/ethnicity (F = 1.079, p = 0.300), nor interaction effect with gender and race/ethnicity (F = 0.593, p = 0.442).

For suicidal ideation, observed differences were not significant in gender (F = 1.584, p = 0.209), race/ethnicity (F = 0.005, p = 0.945), and there was no interaction between them (F = 0.001, p = 0.979). Hence, no significant differences nor interaction effects were observed in suicidal ideation by group.

### Random Forest Classifiers: Model performances

Performances for the random forest classifiers are provided in figures 3-6. Summary score predictions refer to using comorbid scores to predict outcome variables; for instance, summary scores for PHQ-9 and Q9 endorsement were used (along with additional features as listed) to predict a binary outcome for GAD-7, and vice versa.

**Figure 3.**
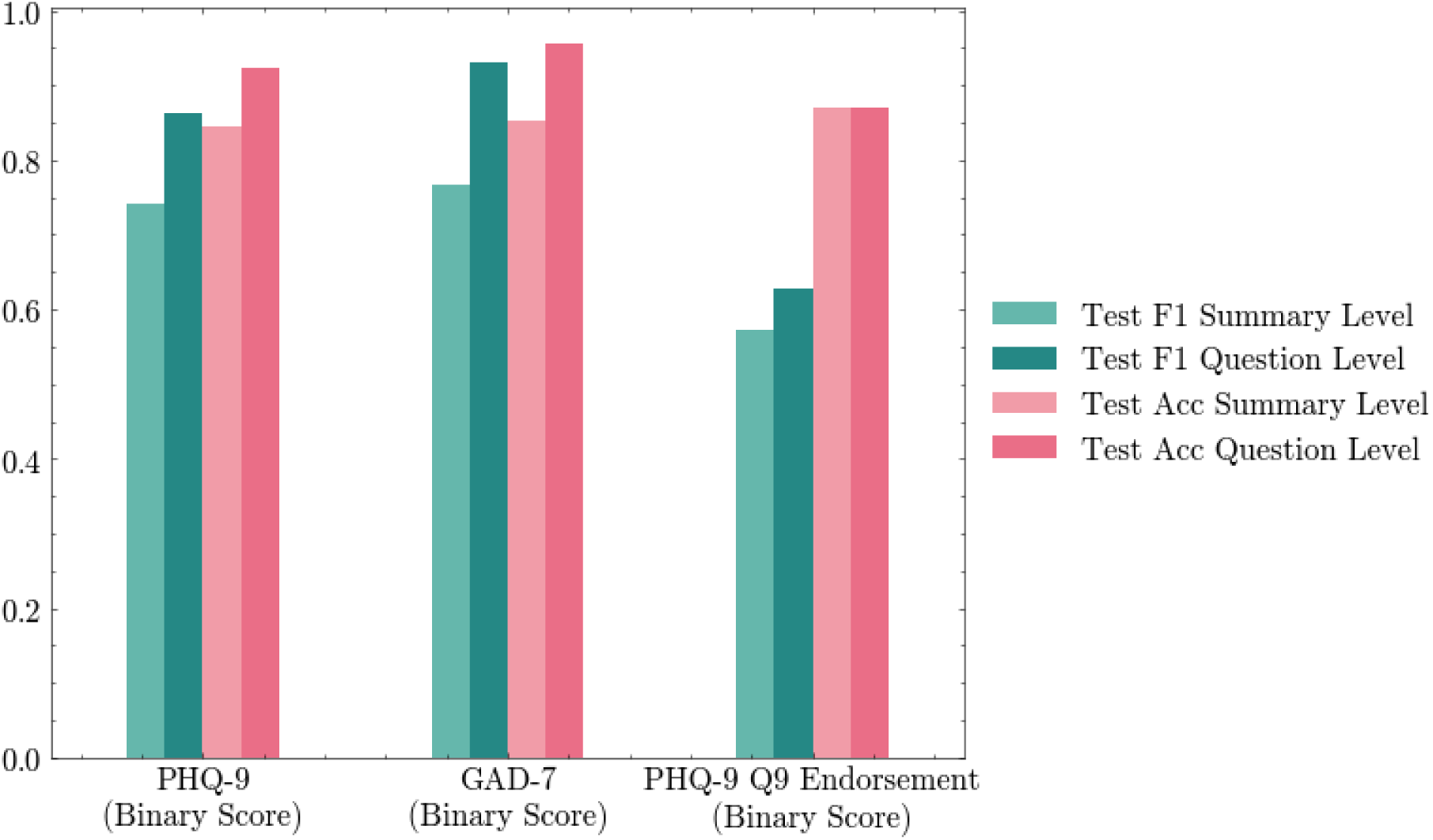
Performance of screening score binary predictions by feature type via RFC

In the overall student population, accuracies for all three predictions (PHQ-9, GAD-7, and Q9 endorsement) were high when using summary or question level data (>0.8). F1 scores were lower in predicting Q9 endorsement (∼0.6). F1 scores were notably lower in predicting suicidal ideation (∼0.6) compared to GAD-7 and PHQ-9 scores (0.7-0.8).

Accuracies for predicting GAD7 score for all groups were high (>0.8, fig. 4). F1 scores for summary score and question data were also in a similar range, with the exception of male and male subgroups, which had F1 scores of approximately 0.5.

**Figure 4.**
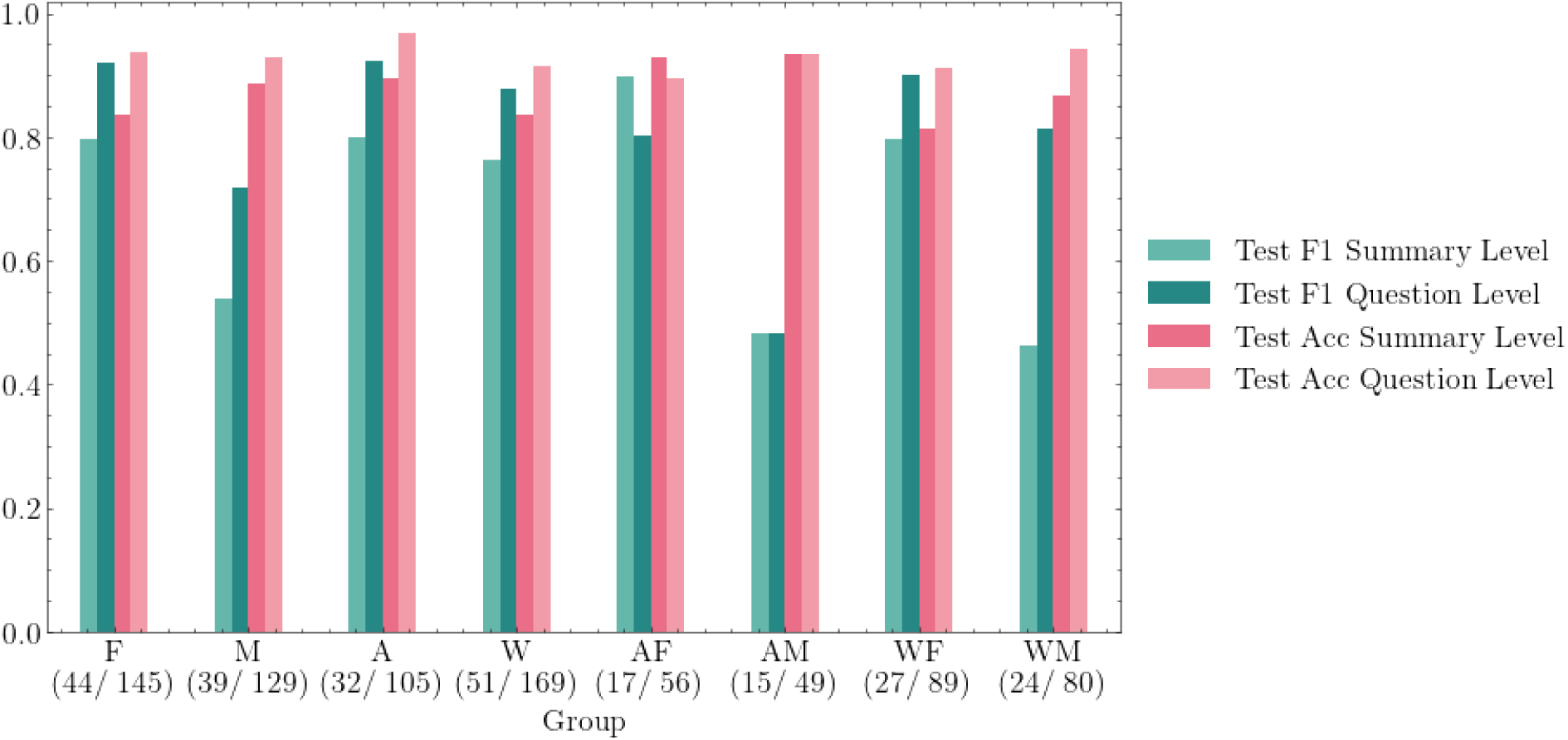
Performance of Clinically Significant Anxiety by student group via RFC. W=White, A=Asian, M=Male, F=Female.

**Figure 5.**
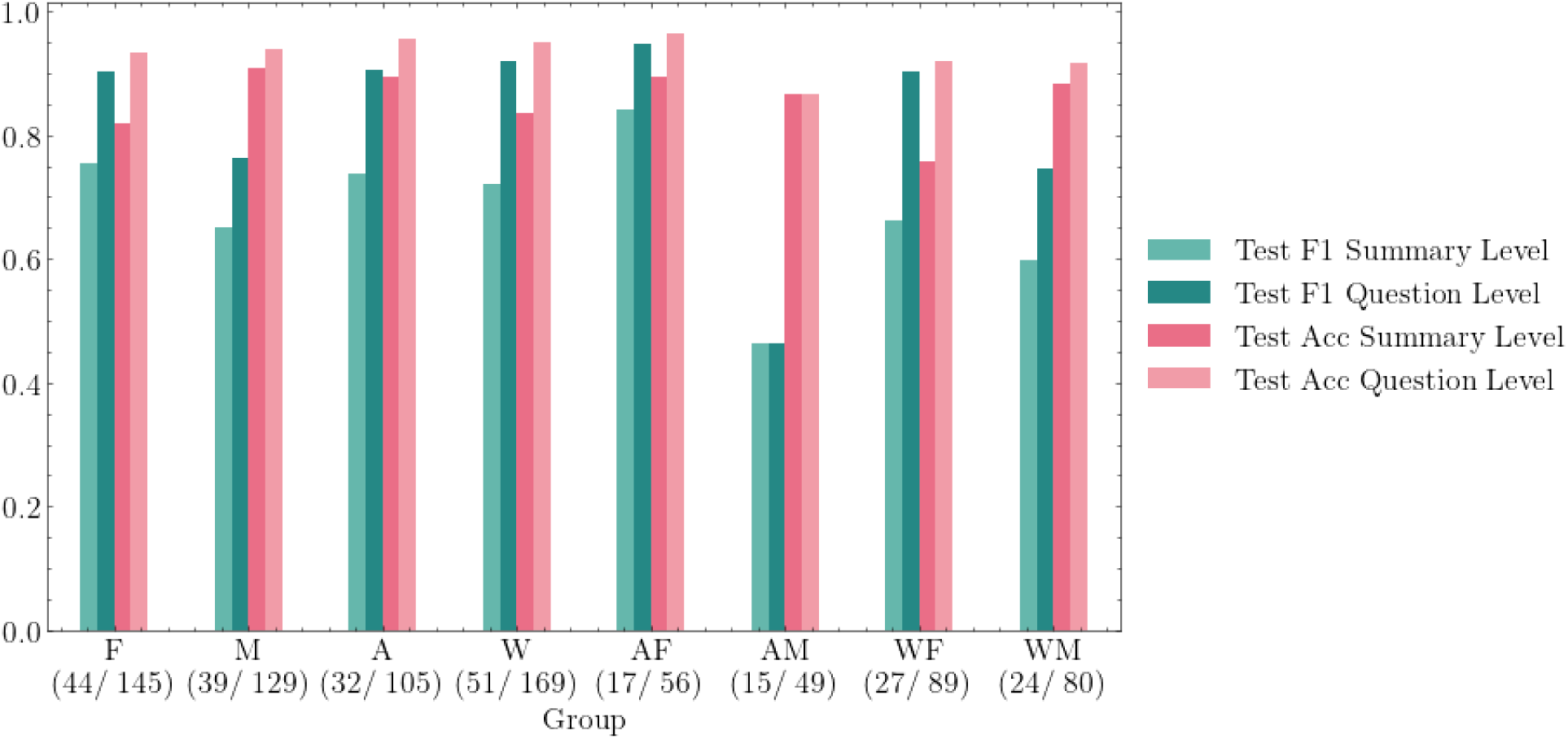
Performance of PHQ-9 score binary predictions by student group via RFC.

**Figure 6.**
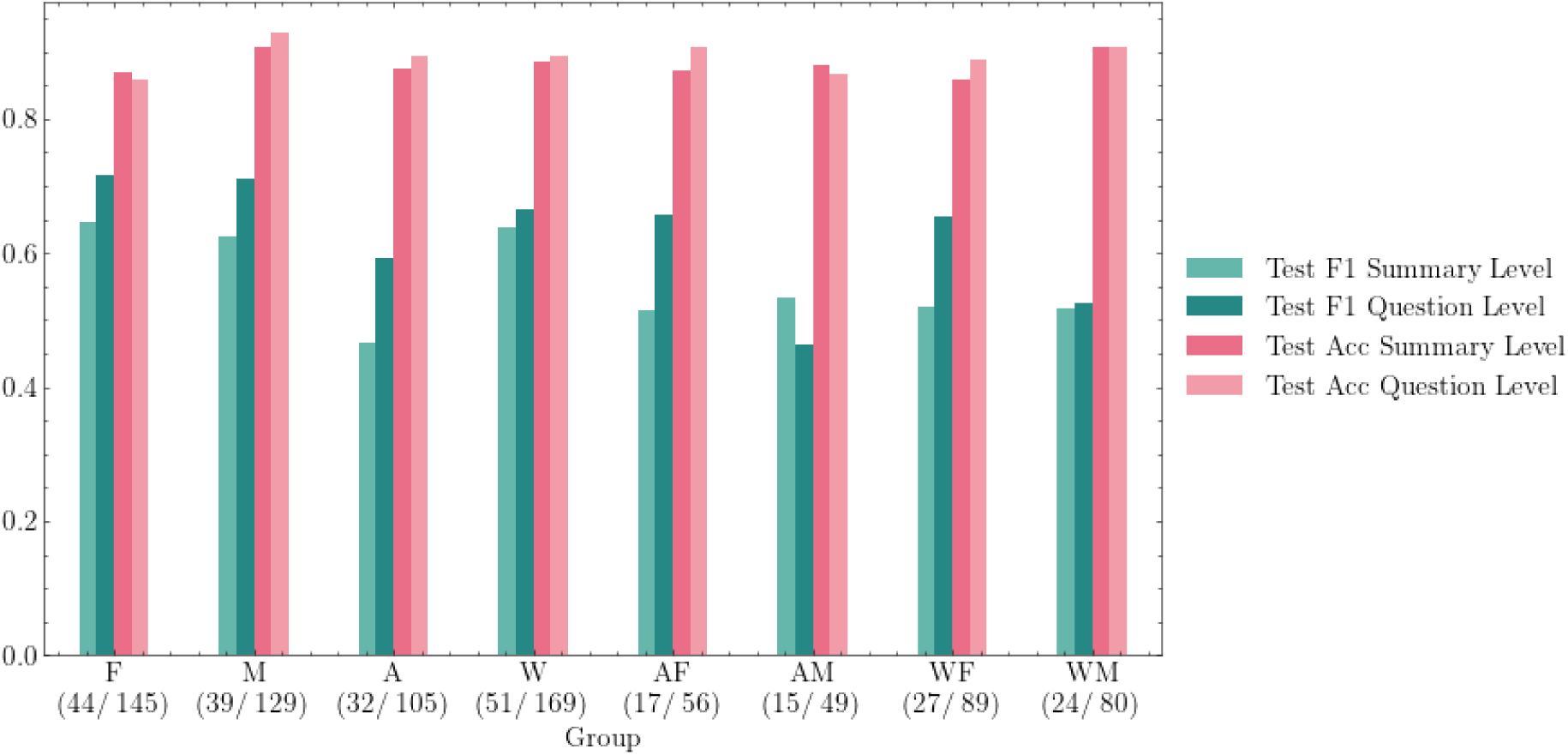
Performance of suicidal risk prediction by student group (PHQ-9 Q9 binary score predictions via RFC). W=White, A=Asian, M=Male, F=Female.

Accuracies for predicting PHQ9 score for all groups were high (>0.8, fig. 5). F1 scores using summary score level data were in the 0.7 - 0.8 range, except lower for the male group, male subgroups, and white female population. However, F1 scores for using question level data were all above 0.8, except in the male group and male subgroups, which were again in the 0.4 - 0.7 range.

Accuracies for predicting PHQ-9 question 9 endorsement for all groups were high (>0.8).

However, F1 scores in all conditions were lower (<0.7).

Overall, the models were highly accurate (85-95%). F1 scores were generally high as well (0.7 to 0.95), notably higher using question level data compared to summary, except in male students and in when predicting suicidal ideation. The low rates can be attributed to the lower numbers of males at higher symptom severities and individuals with suicidal ideation.

### Random Forest Classifiers: Summary Level Feature Contributions

Predictions were performed using comorbid summary scores in addition to student statuses and year end GPA, as in Figures 7-9. In Tables 10-12, the top two average feature contributions across 5 runs are provided. The top two predictors of GAD-7 anxiety severity for the full student population were high PHQ9 scores and low GPA. In most groups, the top predictor was high PHQ-9, however, low GPA was highest in the female subgroups (Asian and White female) (table 7).

**Figure 7.**
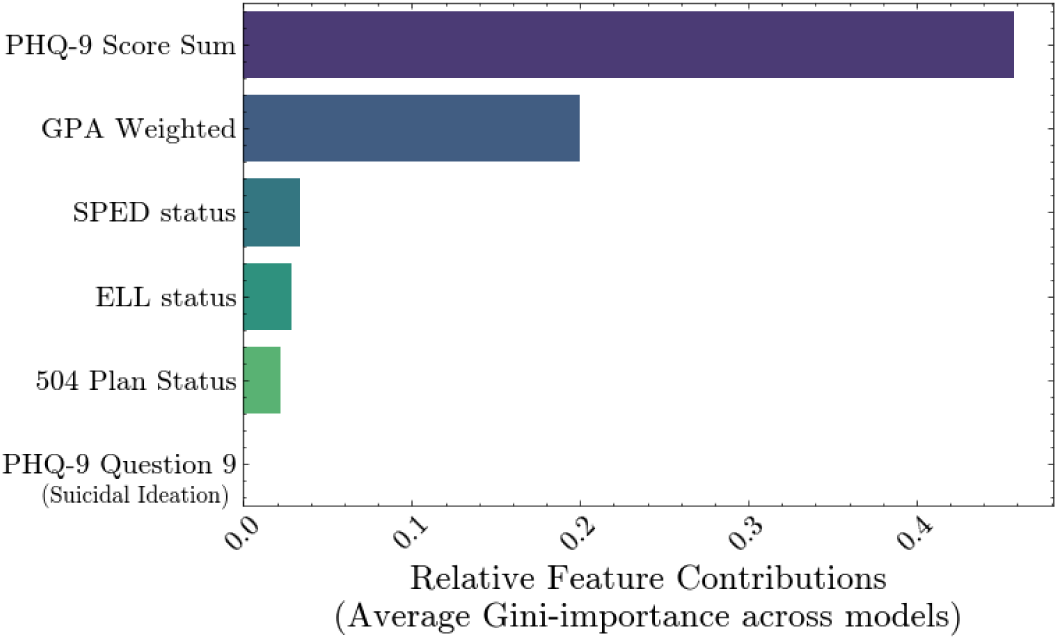
Summary feature ranking for GAD-7 binary score prediction in all students

**Figure 8.**
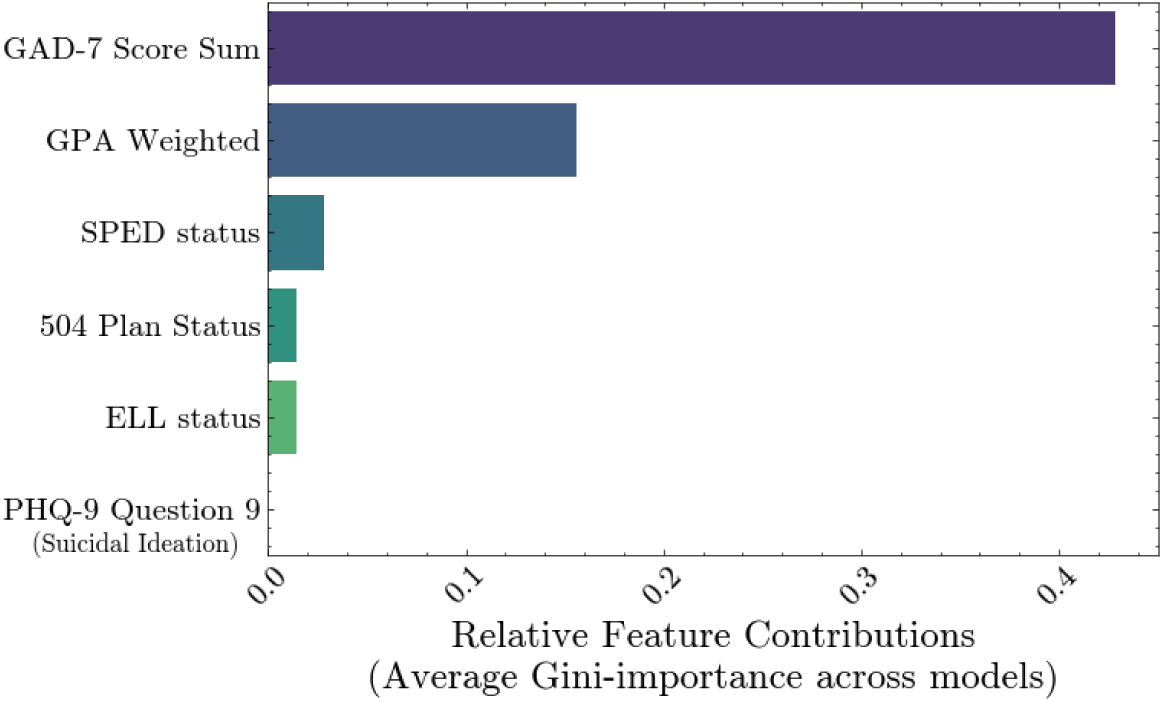
Summary feature ranking for PHQ-9 binary score prediction in all students

**Figure 9.**
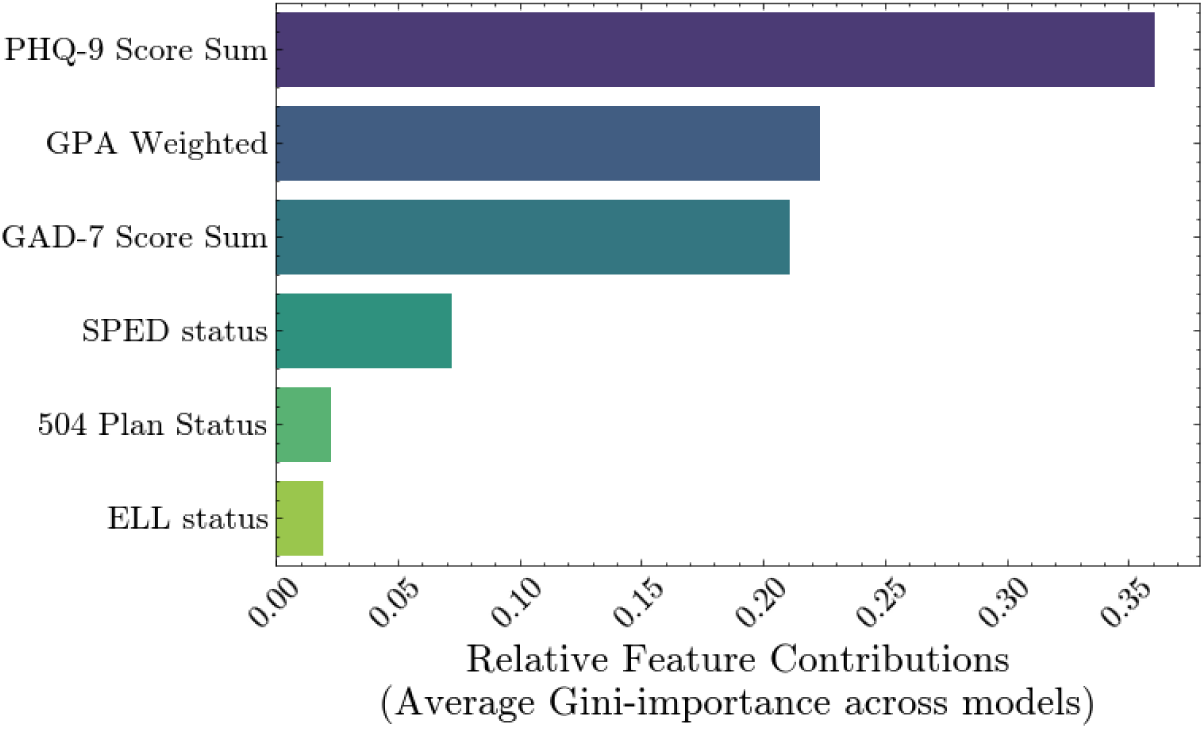
Summary feature ranking for predicting suicidal ideation in all students

**Table 7.** Top 2 questions for GAD-7 binary score prediction by student group.

|  |  | PHQ-9<br>Score | Endorse<br>Q9 | GPA | SPED<br>Status | 504<br>Plan<br>Status | ELL<br>Status |
| --- | --- | --- | --- | --- | --- | --- | --- |
| Gender | F | 1 |  | 2 |  |  |  |
|  | M | 1 |  | 2 |  |  |  |
| Race | A | 1 |  | 2 |  |  |  |
|  | W | 1 |  | 2 |  |  |  |
| Both | AF | 2 |  | 1 |  |  |  |
|  | AM | 1 |  | 2 |  |  |  |
|  | WF | 2 |  | 1 |  |  |  |
|  | WM | 1 |  | 2 |  |  |  |

**Table 8.** Top 2 questions for PHQ-9 binary score prediction by student group. W=White, A=Asian, M=Male, F=Female.

|  |  | GAD-7<br>Score | Endorse<br>Q9 | GPA | SPED<br>Status | 504<br>Plan<br>Status | ELL<br>Status |
| --- | --- | --- | --- | --- | --- | --- | --- |
| Gender | F | 1 |  | 2 |  |  |  |
|  | M | 1 |  | 2 |  |  |  |
| Race | A | 1 |  | 2 |  |  |  |
|  | W | 1 |  | 2 |  |  |  |
| Both | AF | 1 |  | 2 |  |  |  |
|  | AM | 1 |  | 2 |  |  |  |
|  | WF |  |  | 1 |  | 2 |  |
|  | WM | 2 |  | 1 |  |  |  |

**Table 9.** Top 2 questions for PHQ-9 Q9 binary score prediction by student group. W=White, A=Asian, M=Male, F=Female.

|  |  | GAD-7<br>Score | PHQ-9<br>Score | GPA | SPED<br>Status | 504<br>Plan<br>Status | ELL<br>Status |
| --- | --- | --- | --- | --- | --- | --- | --- |
| Gender | F | 2 |  | 1 |  |  |  |
|  | M |  | 2 | 1 |  |  |  |
| Race | A |  | 1 | 2 |  |  |  |
|  | W | 1 | 2 |  |  |  |  |
| Both | AF |  | 1 | 2 |  |  |  |
|  | AM |  | 1 | 2 |  |  |  |
|  | WF |  | 1 | 2 |  |  |  |
|  | WM |  | 2 | 1 |  |  |  |

**Table 10.**
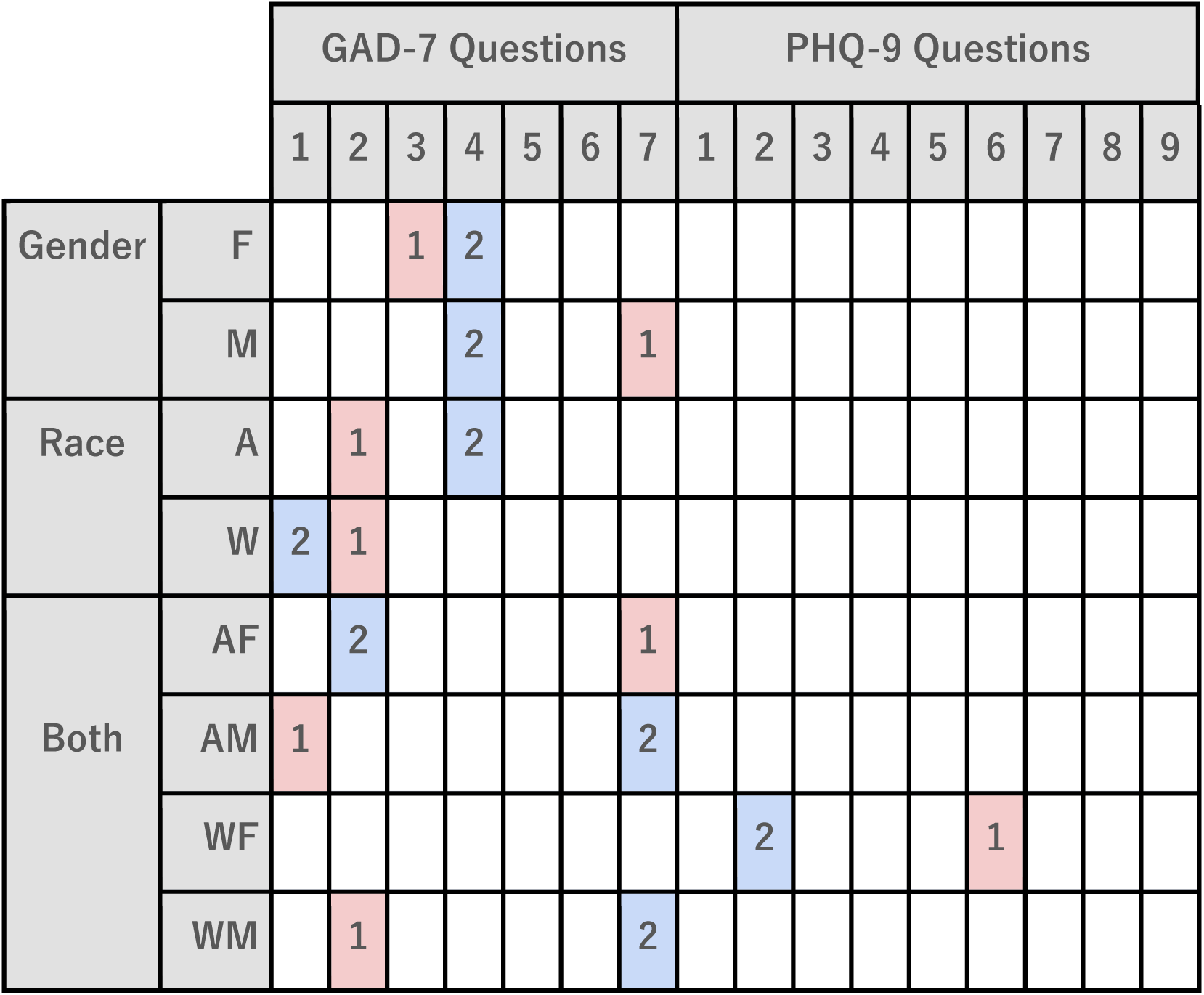
Top 2 questions for GAD-7 binary score prediction by student group. W=White, A=Asian, M=Male, F=Female.

**Table 11.**
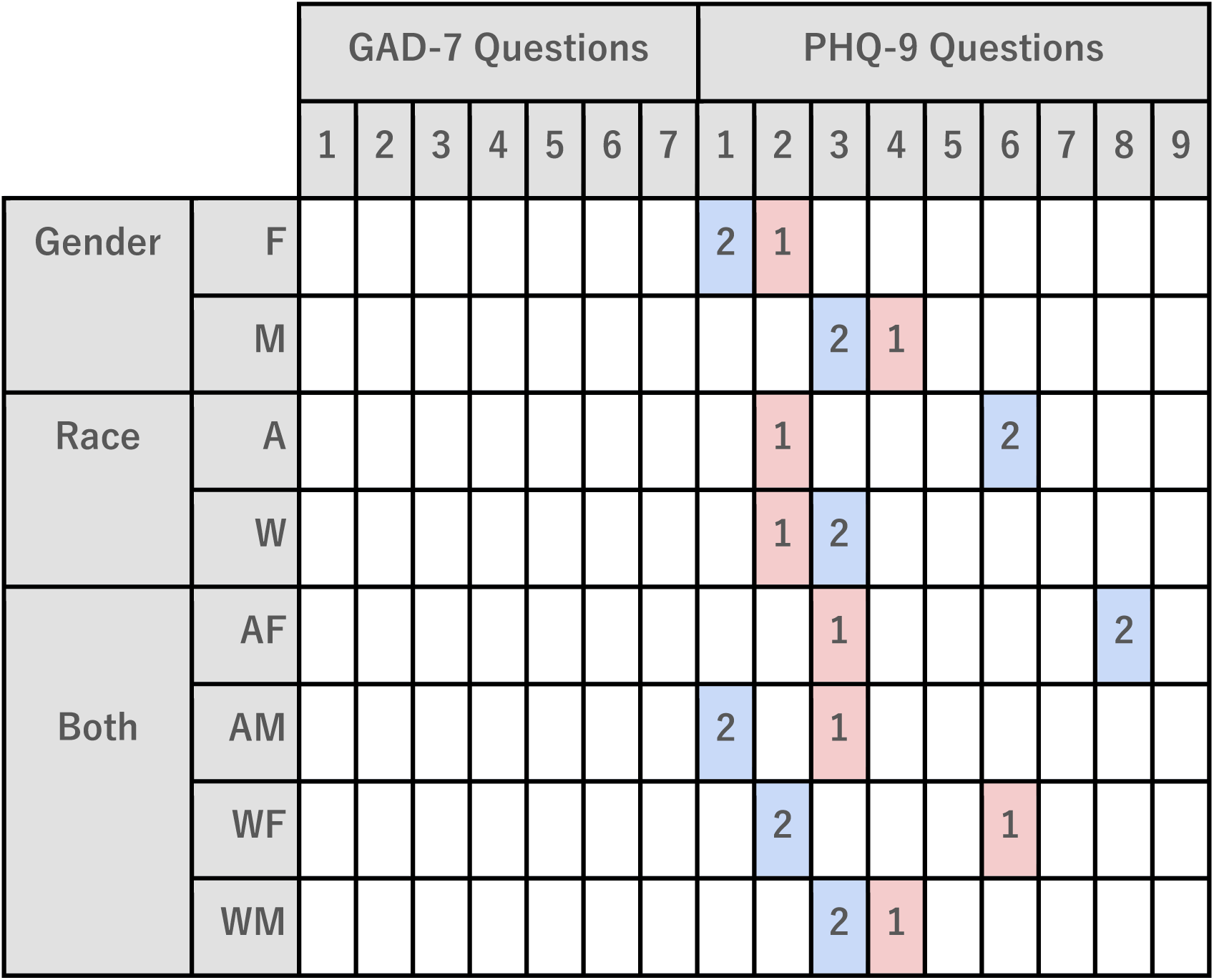
Top 2 questions for PHQ-9 binary score prediction by student group. W=White, A=Asian, M=Male, F=Female.

**Table 12.** Top 2 questions for suicidal ideation prediction by student group.

|  |  | GAD-7 Questions |  |  |  |  |  |  | PHQ-9 Questions |  |  |  |  |  |  |  |
| --- | --- | --- | --- | --- | --- | --- | --- | --- | --- | --- | --- | --- | --- | --- | --- | --- |
|  |  | 1 | 2 | 3 | 4 | 5 | 6 | 7 | 1 | 2 | 3 | 4 | 5 | 6 | 7 | 8 |
| Gender | F |  |  |  |  |  |  |  |  | 1 |  |  |  | 2 |  |  |
|  | M |  |  |  |  |  |  |  | 1 |  |  | 2 |  |  |  |  |
| Race | A |  |  |  |  |  |  |  |  |  |  | 2 |  | 1 |  |  |
|  | W |  |  |  |  |  |  |  | 1 | 2 |  |  |  |  |  |  |
| Both | AF |  |  |  |  |  |  |  |  | 1 |  |  |  | 2 |  |  |
|  | AM |  |  |  |  |  |  |  |  |  |  | 1 |  | 2 |  |  |
|  | WF |  |  |  |  |  |  |  | 2 | 1 |  |  |  |  |  |  |
|  | WM |  |  |  | 2 |  |  |  | 1 |  |  |  |  |  |  |  |

For predicting PHQ-9 score in the full student population, high GAD-7 and low GPA were the top two predictors (fig. 8). This was mostly consistent in all subgroups, except for White females that had GPA and 504 plan status as the top 2 predictors and white males, where low GPA was the top ranked predictor of PHQ-9 score (table 8).

High PHQ-9 score was the greatest predictor for Q9 endorsement in the full student population (fig. 9). The top two feature contributions for student groups were most often PHQ-9 score and low GPA, with White students having high GAD-7 as a stronger predictor (table 9). GPA was the top ranked feature, outranking high PHQ-9 score in the female, male, and White male subgroups.

Overall, for summary score level predictions in the total student population, a comorbid summary score was the highest predictor; GAD-7 for PHQ-9 prediction, PHQ-9 for GAD-7 prediction and PHQ-9 for Q9 endorsement prediction. GPA was ranked 2nd in all three predictions as well. For group specific predictions, most groups followed suit, with a few exceptions. For GAD-7, both female subgroups had GPA higher than PHQ-9. For PHQ-9, the White female population had 504 plan status as the 2nd top feature. For Q9 endorsement, the White population had GAD-7 score as a higher ranked predictor than PHQ-9 score. This suggests anxiety to be a higher predictor for suicidal ideation in the White population compared to depressive symptoms.

### Random Forest Classifiers: Question Level Feature Contributions

For the question level analysis, predictions of GAD-7 and PHQ-9 scores and Q9 endorsement used responses to the GAD-7 and PHQ-9 questions as features; both question types were provided to assess for any predictive crossover. The top features for the predictions on the overall population are shown in figures 10-12. Tables 10-12 display the top two features when predicting for each student group.

**Figure 10.**
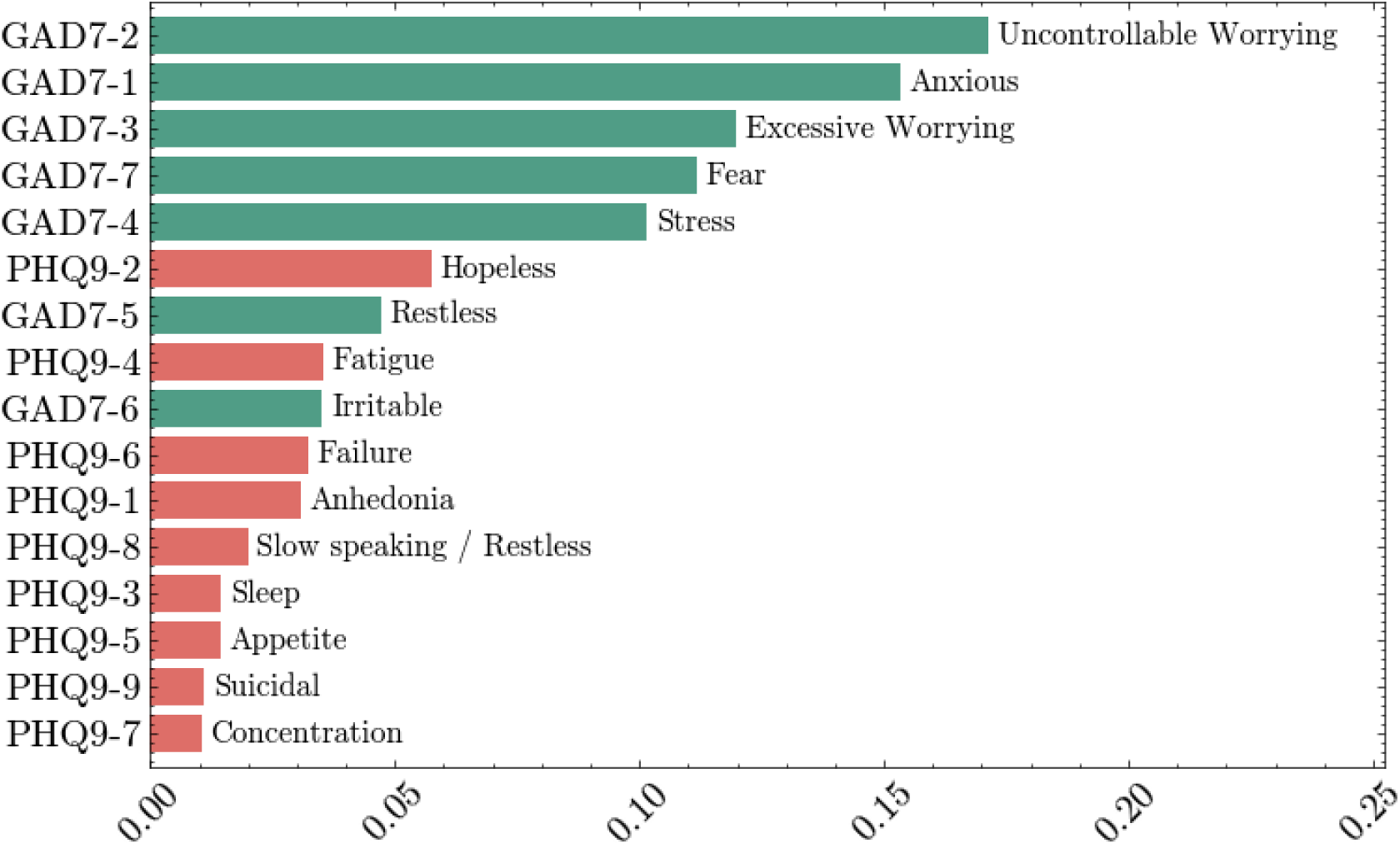
Question feature ranking for GAD-7 binary score prediction in all students

For all students, questions associated with worrying and anxiousness ranked highest in the prediction of GAD-7 score (fig. 10). However, across individual student groups, this varied significantly (table 10). By gender, worrying ranked highest for female students and fearfulness was top ranked in males. For White female students, PHQ-9 questions ranked as the highest predictors of anxiety (hopelessness and failure). Overall, most groups had GAD-7 questions concerning worrying, stress, and fear in their top 2 contributions.

PHQ-9 questions related to anhedonia, hopelessness, and sleep were most predictive of PHQ-9 score in all students (fig. 11). As with GAD-7 prediction, the top 2 ranked features varied substantially between groups (table 11). There was no overlap by gender alone; for females, this was hopelessness & anhedonia, and for males, fatigue and sleep difficulties. Fear of failure ranked second for Asian students and slow speaking/restless ranked second for Asian females. This suggests distinct differences in their experiences of depression.

**Figure 11.**
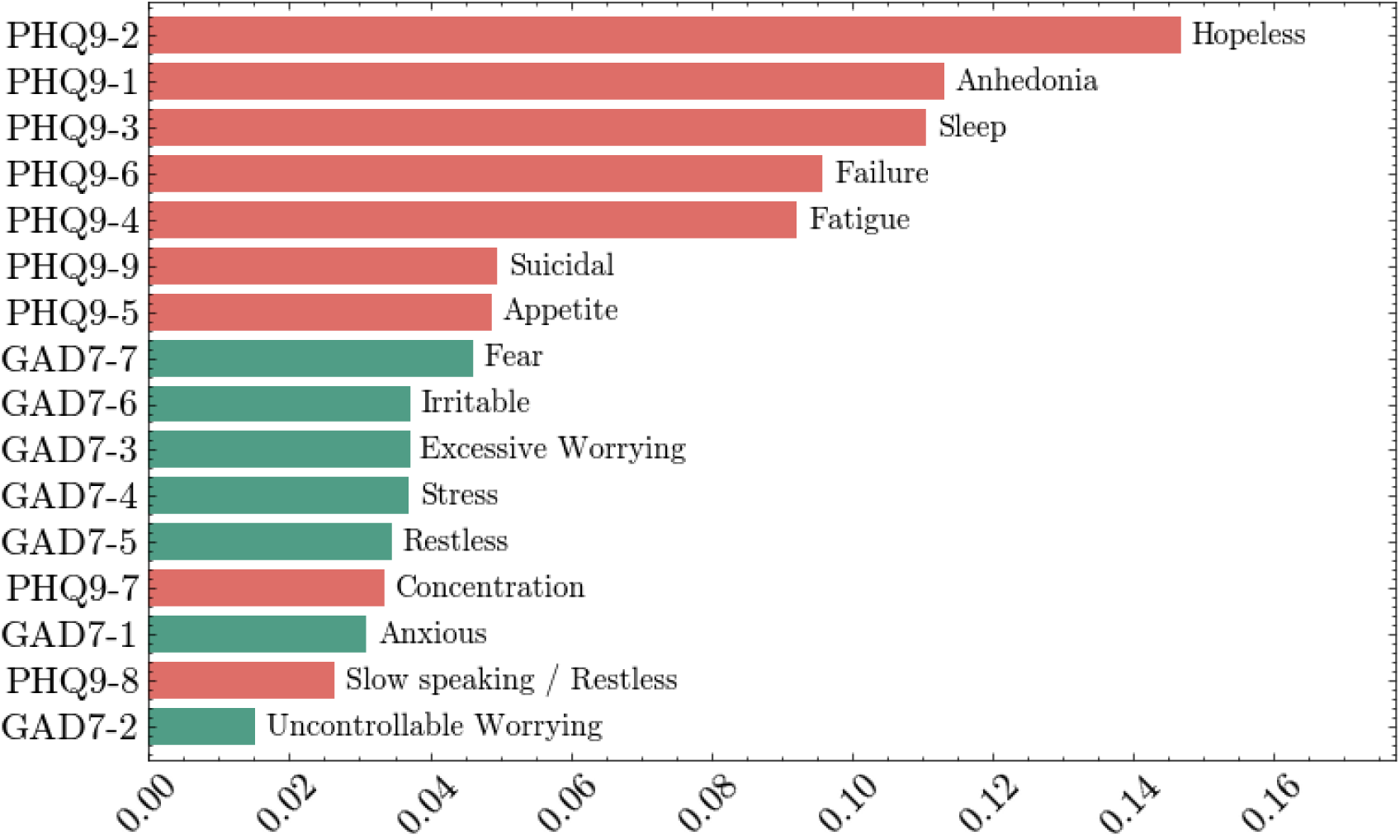
Question feature ranking for PHQ-9 binary score prediction in all students

**Figure 12.**
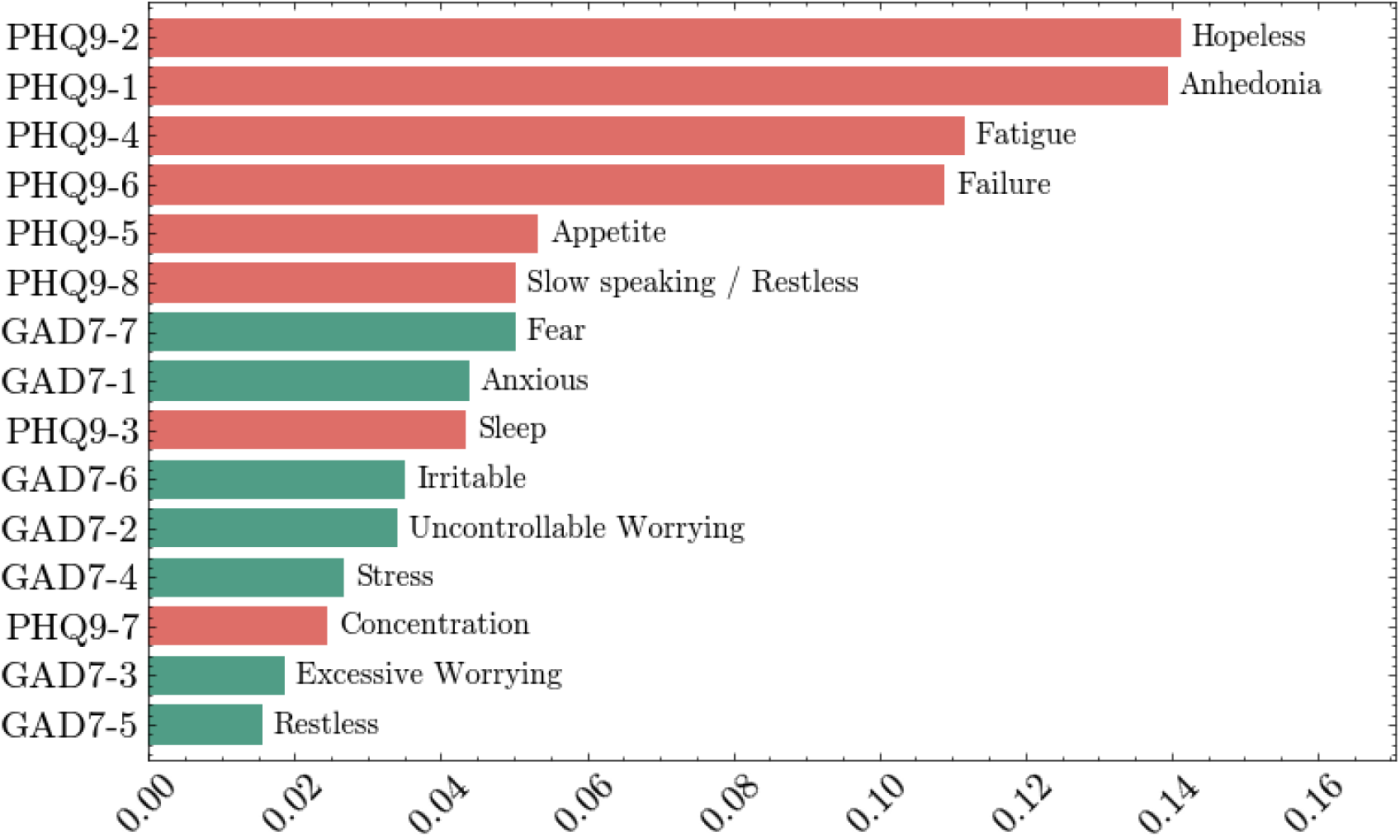
Question feature ranking for suicidal ideation prediction in all students

For the entire student population, hopelessness and anhedonia ranked the highest in predicting suicidal ideation, followed by fatigue and failure (fig. 12). These top two features were consistent across several groups, however, there was still notable variation (table 12). For instance, in females, hopelessness and failure were highest whereas anhedonia and fatigue were highest in males. In Asian students, failure and fatigue related questions were ranked highest, whereas anhedonia and hopelessness questions were most predictive in White students. Within males, Asian males had fatigue and failure ranked highest, while White males had anhedonia and trouble relaxing (Q4 from GAD-7, the only GAD-7 question ranked in the top two).

## Discussion

The purpose of this paper was to identify high school students with greater depression severity, anxiety severity, and who reported suicidal ideation. This study categorized Asian, White, male, and female students with clinically significant anxiety, depression, and with suicidal ideation, from one high school in Massachusetts. Our primary aim with this study was to predict mental health outcomes using student demographic, academic, and detailed clinical mental health screening data. Since there are substantial differences in prevalence rates across gender and racial and ethnic groups, we geared our analysis towards predicting anxiety, depression, and suicidal ideation by student gender and race groups. Our highly accurate analysis identified an unexpectedly strongly predictive inverse relationship between GPA and depression and anxiety screening scores. We also report substantial group-specific variation in predictors of screening scores, highlighting the importance of considering individual and group specific targeting and intersectionality in mental health research, programing, prevention, and treatment.

### Summary level findings

Overall, even with a modest dataset, clinical outcomes could be predicted with high accuracy using Random Forest with summary mental health screener and student data. While the F1 scores were generally high as well, they were substantially lower in male groups and with predicting suicidal ideation. This is likely due in part to smaller samples sizes, but could also be related to decreased reporting in males.

Substantial group differences were observed in the summary score predictions. The high performances of the models in predicting anxiety severity scores with depression severity scores, and vice versa, underscores the comorbidity of these disorders in students. Although the comorbidity of depression and anxiety is established, the present results indicate that comorbid symptoms are stronger predictors in some groups. Concerning suicidal ideation, predictions of Q9 endorsement reveal that GAD-7 score ranks higher than PHQ-9 for White females, suggesting the importance of considering anxiety in the development of targeted interventions for suicidal ideation for this population. This aligns with previous reports of increased prevalence of anxiety in female populations (Källström et al., 2022; McLean et al., 2011; Pigott, 1999). Another (Keogh et al., 2004)year-end GPA, where low GPA predicted higher anxiety and depression scores and suicidal ideation.

Lower academic performance has been associated with depression and anxiety levels in adolescents (Owens et al., 2012) and the work of Aronen et al.(Aronen et al., 2005) has suggested the need to holistically consider anxious and depressive traits as they relate to cognition and academic performance. Academic performance is known to be a significant source of stress in high school students (Barbayannis et al., 2022; Pascoe et al., 2020; Suresh, 2015). Selective attention and concentration are increased when academic stress is decreased (Fernández-Castillo and Caurcel, 2015). The present analysis revealed GPA as an unexpectedly high ranking (greater than depression score) predictor of anxiety female students and White males. The relationship between anxiety and academic performance has been explored (Aronen et al., 2005; Christensen, 1979; Keogh et al., 2004) and both trait and state-anxiety lead to academic performance deficits, mediated through working memory (WM) inhibition (Maloney et al., 2014). Individuals with higher levels of math anxiety are 20% more likely to make mistakes in conditions with high WM requirements compared to individuals with lower levels of math anxiety. In addition to the findings on anxiety and depression, GPA was the highest ranked feature for predicting suicidal ideation in both female and male groups, where it was driven by data from White males. In discussions with the school district staff, it was expected that there would be a positive relationship between GPA and depression and anxiety scores due to patterns in treatment seeking; the opposite of what was observed. This finding may be valuable to the vast majority of school districts that do not conduct clinical mental health screening. The predictive strength of the negative relationship between GPA and anxiety, depression, and suicidal ideation indicate the need for a greater appreciation of the relationship between the two.

While there may be effective interaction between academics and mental health resources in some school districts, a more integrated approach in the form of enhanced coordination and communication could provide benefit to both dimensions. For instance, programming and interventions on the adverse effects of anxiety on GPA may have mutual benefits for both academics and mental health. Punitive or disciplinary responses to low GPA, such as restricting participation in sports that have beneficial effects on mental health (Appelqvist-Schmidlechner et al., 2018; Easterlin et al., 2019), may be counter-productive and have adverse effects compared to a more mental health informed response. A greater awareness and understanding of the strength and impact of the relationship between academic performance and mental health, especially with regards to suicidal ideation, in students, parents, faculty, and staff may lead to decreased stigma and earlier and more effective prevention and intervention.

### Question level findings

In comparison to the summary level findings, the screening question level data provided a more nuanced, symptom specific view of the variability in student depression and anxiety. For instance, analysis of all students did not identify fearfulness as a high ranking predictor despite it being a key predictor of anxiety in males. This issue with missing key predictors in the male population is systematic due to higher PHQ-9 and GAD-7 scores in the female population. This created a masking effect. In PHQ-9 prediction, hopelessness and anhedonia were the high ranked predictors in the overall population and females, while for males, fatigue and sleep were ranked highest. Furthermore, even within the female population, Asian females had sleep difficulties and slow speech/restlessness as a greater predictor of PHQ-9 scores. Thus, recommendations decision making based on the whole population may align with majority subgroups (such as females in general), while masking the needs/concerns of minority subgroups (such as males and Asian females). This is a concerning phenomenon given the higher rates of suicides in males compared to females (Stone and Report, 2023).

In male students, who may be more reluctant to disclose mental health challenges to teachers and healthcare professionals than female students (Chatmon, 2020; Clark et al., 2020; McKenzie et al., 2022), our findings indicate that fatigue and sleep difficulties are crucial factors to consider when assessing their mental well-being. Sleeping less than 7 hours on weekdays or weekends is associated with poorer performance in school (Ming et al., 2011), and several other studies have found significant detrimental effects of poor sleep quality on academic performance (Ahrberg et al., 2012; Kang et al., 2012; Shin et al., 2003). Factors such as skipping class, school achievement, disliking school, and pressure from schoolwork contribute to student fatigue (Matos et al., 2016).

Furthermore, fatigue and sleep may be causal and relatively more actionable within home and school settings, such as implementing earlier bedtimes, reducing screen time, removing screens from bedrooms in the evening, and altered school start times (Åslund et al., 2018; Busch et al., 2017; Gadam et al., 2023). Depression and sleep disturbances have a bidirectional relationship, where insomnia is known and modifiable risk factor for depression (Alvaro et al., 2017; Roane and Taylor, 2008; Roberts and Duong, 2013; Soltani et al., 2023).

Treatment of insomnia has the potential to prevent subsequent depression. Screening tests that assess sleep disturbance related challenges may facilitate the early identification of sleep disturbance related subtypes of depression and guide prevention, early intervention, and treatment. An increased understanding of adolescents’ sleep needs has already prompted many school districts across the US to adopt later start times (Kelman, 1999; Malone, 2011). School districts that still have relatively early high school start times may see improvements in academic performance and mental health from later start times and/or fatigue and sleep related programming, education, and individual intervention.

Group variability in mental health symptoms is also highlighted by PHQ-9 questions being strong predictors of anxiety in the White female population. This implies that depression-specific interventions for could be an essential consideration for treating anxiety in some students, and that comprehensively addressing the anxiety/depression comorbidity is crucial.

These findings indicate the importance of evaluating different metrics when assessing mental health among diverse student populations, allowing for more personalized and effective support and interventions.

In the present female population, the consistently robust role of hopelessness and anhedonia may be associated with social media consumption. The social media habits between genders have been explored in the literature. Social media is higher in both perceived usefulness and perceived risk in females than in males. Females are more likely to compare themselves with other females, while males used social media to connect (Haferkamp et al., 2012; Towner et al., 2022). A study of 221,096 adolescents, 13 to 18-years-old, in the U.S. and UK found adolescent girls to spend more time on social media, texting, and smartphones (Twenge and Martin, 2020). They also found increased technology use to be twice as likely to be associated with reduced well-being, increased mental health issues, and suicidal risk in females compared to males. More recent reviews of studies investigating depression and/or anxiety and social media use in youth populations have identified strong bidirectional relationships that differ by gender (Azem et al., 2023; Lopes et al., 2022). These relationships are likely driven by problematic social media use (Cunningham et al., 2021).

These previous findings underscore the crucial consideration of gender differences in social media use and/or effects and its potential impact on mental health, particularly among adolescent girls. Strategies to promote healthy and balanced use of technology should be considered. While anhedonia is a general and nonspecific symptom that may not be directly influenced by the inherent rewarding aspects of a student’s activities, it is possible that reevaluating and modifying the selection, scheduling, and management of academic and non-academic pursuits to decrease stress and maximize reward could lead to a reduction in hopelessness, anhedonia, and overall depression levels. By addressing the potential impact of social media and other lifestyle factors on specific symptoms such as hopelessness and anhedonia, interventions can be tailored to support the mental well-being of female students.

Though overall predictors are applicable to all groups to some degree, it may be more effective to identify intervention strategies that specifically address the most pressing issues faced by different student populations to efficiently and effectively allocate effort and resources and optimize treatment outcomes. Additionally, evaluation of student mental health globally may overshadow critical group specific needs, decreasing efficiency and effectiveness in identifying and treating these at-risk groups. For students with unique symptoms that are not being addressed, this could create an adverse attitude/response towards efforts to support mental health. Differences in predictive ranking support the need for personalized recommendations, data collection, and care for the individual student. Enhanced communication and coordination among members of specific support structures, such as academics and counseling, may lead to synergistic improvements in both mental health and academics. Beyond the scope of this project, the predictive importance of non-academic factors such as disciplinary actions, social interaction (withdrawal, aggression) and impulsivity should also be considered as key risk factors for anxiety, depression, and suicidal ideation.

Despite the limited descriptive statistics on the present student population, our analyses provided several unbiased patterns in gender and race. Analogous to the benefits of teachers getting to know students in an academic setting, detailed data collection can improve mental health programming, prevention, targeting, and interventions. As physical examinations are required for scholastic sports participation and standardized exams for graduation, standardized mental health data collection or clinical mental health screening is warranted given the concerning increase in mental illness prevalence among children and adolescents.

### Limitations

There were several limitations to the present study. Firstly, the numbers of individuals in some groups and with some features were low, which restricted the depth of our analysis and model performance. Due to the demographics of the school district, some student populations had less than 10 individuals (gender non-conforming, Black, Latinx, and Hispanic) and could not be analyzed. A targeted survey of underrepresented individuals or school districts with greater representation of such students is warranted. It should be noted that the overall F1 scores in both the summary level and question level predictions, stratified and unstratified, were relatively lower compared to the high accuracies. Despite this, F1 and accuracy scores were generally high, suggesting the robustness of RFC and their ability to predict mental health outcomes in student groups in a targeted way.

The feature set provided was restricted to a small set of variables that likely did not capture the comprehensive complexity of mental health experiences. Additionally, mental health measures were not taken continuously, so we were unable to track how mental health changed over time. Due to the way the data were collected, we were making predictions on past mental health based on future GPA data,.

## Conclusion

In conclusion, our study has emphasized the importance of considering academic performance and intersectionality in mental health research and treatment among student populations. By utilizing machine learning tools, we were able to identify unique patterns in mental health symptoms and predictors across different student groups. We found that anxiety and depression can manifest differently across student gender and racial groups. This underscores the need for personalized data collection recommendations, and care for individual students. Enhanced communication and coordination among members of specific social spheres, through academics and counseling, can help address these unique needs.

## Data Availability

All data in the present study were provided to WPI through a data use agreement.

## Notes

### Competing Interest Statement

The authors have declared no competing interest.

### Funding Statement

This study did not receive any funding.

### Author Declarations

The Worcester Polytechnic Institute IRB waived ethical approval for this work.

